# Sensitivity, specificity and avoidable workload of using a large language models for title and abstract screening in systematic reviews and meta-analyses

**DOI:** 10.1101/2023.12.15.23300018

**Authors:** Viet-Thi Tran, Gerald Gartlehner, Sally Yaacoub, Isabelle Boutron, Lukas Schwingshackl, Julia Stadelmaier, Isolde Sommer, Farzaneh Aboulayeh, Sivem Afach, Joerg Meerpohl, Philippe Ravaud

**Affiliations:** Université Paris Cité, CRESS, INSERM, INRAE, Paris, France; Centre d’Epidémiologie Clinique, Hôpital Hôtel-Dieu, AP-HP, Paris, France; Department for Evidence-based Medicine and Evaluation, University for Continuing Education Krems, Krems, Austria; Center for Public Health Methods, RTI International, Research Triangle Park, USA; Institute for Evidence in Medicine, Medical Center - University of Freiburg, Faculty of Medicine, University of Freiburg, Freiburg, Germany; Epidemiology in Dermatology and Evaluation of Therapeutics (EpiDermE) - EA 7379, University Paris Est; Department of Epidemiology, Columbia University Mailman School of Public Health, New York, NY, USA

**Keywords:** Systematic reviews, generative artificial intelligence, methodology

## Abstract

**Importance:** Systematic reviews are time-consuming and are still performed predominately manually by researchers despite the exponential growth of scientific literature.

**Objective:** To investigate the sensitivity, specificity and estimate the avoidable workload when using an AI-based large language model (LLM) (Generative Pre-trained Transformer [GPT] version 3.5-Turbo from OpenAI) to perform title and abstract screening in systematic reviews.

**Data Sources:** Unannotated bibliographic databases from five systematic reviews conducted by researchers from Cochrane Austria, Germany and France, all published after January 2022 and hence not in the training data set from GPT 3.5-Turbo.

**Design:** We developed a set of prompts for GPT models aimed at mimicking the process of title and abstract screening by human researchers. We compared recommendations from LLM to rule out citations based on title and abstract with decisions from authors, with a systematic reappraisal of all discrepancies between LLM and their original decisions. We used bivariate models for meta-analyses of diagnostic accuracy to estimate pooled estimates of sensitivity and specificity. We performed a simulation to assess the avoidable workload from limiting human screening on title and abstract to citations which were not “ruled out” by the LLM in a random sample of 100 systematic reviews published between 01/07/2022 and 31/12/2022. We extrapolated estimates of avoidable workload for health-related systematic reviews assessing therapeutic interventions in humans published per year.

**Results:** Performance of GPT models was tested across 22,666 citations. Pooled estimates of sensitivity and specificity were 97.1% (95%CI 89.6% to 99.2%) and 37.7%, (95%CI 18.4% to 61.9%), respectively. In 2022, we estimated the workload of title and abstract screening for systematic reviews to range from 211,013 to 422,025 person-hours. Limiting human screening to citations which were not “ruled out” by GPT models could reduce workload by 65% and save up from 106,268 to 276,053-person work hours (i.e.,66 to 172-person years of work), every year.

**Conclusions and Relevance:** AI systems based on large language models provide highly sensitive and moderately specific recommendations to rule out citations during title and abstract screening in systematic reviews. Their use to “triage” citations before human assessment could reduce the workload of evidence synthesis.

## 1. Introduction

Evidence synthesis is defined as the “*process of bringing together information and knowledge from many sources to inform decisions*” ^1^. According to prominent researchers, it is considered as one of the most valuable contributions research can offer decision-makers ^1,2^. Yet, comprehensive evidence synthesis in the form of high quality systematic reviews, with or without meta-analysis, is limited by the fact that researchers still often manually screen thousands of studies to determine whether they meet the eligibility criteria of reviews, despite the exponential growth of scientific literature ^3^. Manual screening is time-consuming because only a limited fraction (often less than 10%) of the screened studies is finally included. Worse, this screening process is conducted in duplicate because of the risk of error (with a 7 to 11% error rate during abstract screening) ^4-6^. A review estimated that conducting a single systematic review required over a thousand hours of highly skilled manual labour, with hours increasing or decreasing proportionally to the body of evidence to be screened ^7^.

Among methods to increase the efficiency of screening for evidence synthesis, automated tools using natural language processing and/or machine learning methods have been developed. Most tools work by prioritizing relevant studies learning from the researchers’ decision to include the study or not, via active “human-in-the-loop” learning ^8-10^. In a survey study, such tools provided on average 40% of time savings as compared to manual screening. Yet, these tools have limitations. For example, their performance depends on the proportion of relevant publications (i.e., publications that will be included) and the complexity of the inclusion criteria used by the research team ^11,12^.

Recently, the development of general-purpose Artificial Intelligence (AI) systems based on large language models (LLMs), such as ChatGPT (OpenAI), has changed the paradigm of task automation. LLMs have shown excellent ability in answering health questions ^13,14^, diagnosing conditions ^15^, and performing at (or near) the passing threshold for the exams of the United States Medical Licensing Exam (USMLE) ^16^, without any specialized training or reinforcement. LLMs can also accurately label unannotated data (accuracy 89% vs. 95% for human labelling) ^17,18^. Three studies have investigated the performance of LLMs to screen for systematic reviews in different fields (informatics and literature) with sensitivities ranging from 60% to 90% and specificities ranging from 10 to 60% depending on the dataset examined ^19-21^. Beyond their performance, LLMs also transform the accessibility of automated tools for non-specialists thanks to the use of chatbot interfaces where users can instruct the models in natural language.

In this study, we aimed to appraise how a LLM-based system could perform title and abstract screening in systematic reviews. We 1) developed a set of prompts for the Generative Pre-trained Transformer (GPT) 3.5-Turbo model (OpenAI) aimed at mimicking the process of title and abstract screening by human researchers; 2) evaluated the performance of the developed prompts in five high quality systematic reviews; and 3) performed a simulation study to estimate the avoidable workload when using LLMs to screen citations for inclusion in systematic reviews.

## 2. Methods

We investigated the sensitivity, specificity and avoidable workload when using GPT 3.5-Turbo models for screening in systematic reviews.

### 2.1. Data sources

Unannotated bibliographic databases from five systematic reviews conducted by researchers from Cochrane Austria, Germany and France were used in this study: 1) a review on the efficacy of primary treatment of confirmed COVID-19 in outpatient settings (Sommer et al., 2022) ^22^; 2) a review on the efficacy of outpatient treatment options for the omicron variant of SARS-CoV-2 (Sommer et al., 2023) ^23^; 3) a methodological review on the epidemiology and reporting characteristics of non-randomized studies of pharmacologic treatment (unpublished); 4) a review on the effects of dairy intake on intermediate disease markers in adults (Kiesswetter et al., 2023) ^24^; and 5) a Cochrane review on systemic pharmacological treatments for chronic plaque psoriasis (Sbidian et al., 2023) ^25^. In all systematic reviews, decisions to include a citation beyond title and abstract screening were performed by two reviewers in duplicate and independently. All systematic reviews were published after January 2022 and hence were not in the training data set from GPT 3.5-Turbo.

Unannotated bibliographic databases contained the deduplicated list of citations (i.e., studies identified by their title and abstracts) retrieved from electronic search, the list of citations selected after examination of title and abstracts only, and the list of citations selected based on their full texts. In addition to the bibliographic databases, the registered the protocols were used for the inclusion and exclusion criteria of studies in the reviews (PROSPERO CRD42022323440, CRD42023406456, and CRD42022303198, OSF https://osf.io/ywr8s/, and https://doi.org/10.1002/14651858.CD011535).

### 2.2. Prompt development

One investigator (VTT) developed five prompts for the GPT 3.5-Turbo model aimed at mimicking how humans apply the PICOS framework (Population, Intervention, Control, Outcomes and Study design) during title and abstract screening. Each prompt was focused on one element of the PICOS framework and instructed the model to 1) extract relevant information from the title and abstract of citations retrieved from electronic searches and; 2) assess whether the extracted information corresponded to the criteria reported in the protocol of the systematic review (using the same words as those used in the protocol); and 3) give a recommendation to “include” or “exclude” the citation ^26^. For example: *“Read the following text and pay close attention to details. Proceed step by step. First, assess the control. Second, answer with the words “YES” or “NO” by using the following algorithm: 1-If the control is a placebo, or usual care, or a different dose or duration of the intervention, answer “YES”. 2-If the control is an active treatment, different from the intervention, answer “NO”. 3-If unclear, answer “NO”.”* We used temperature hyperparameters of “0”, so as to have deterministic outputs. Final prompts for each review are detailed in **Appendix 1**.

The investigator (VTT) designed a program applying the series of prompt to each citation so as to sequentially assess the design, population, intervention, control and then outcomes. The program was instructed to stop whenever the title or abstract did not fit the inclusion and exclusion criteria of the review. Both prompts and the program were developed with 10 to 20% of citations by comparing the output from the model to the decisions from the authors of systematic reviews and optimizing the ability of the model to “rule out” citations. Final prompts were run on all citations through the application programming interface (API) of GPT. Final output for each review was a binary recommendation (Yes/No) to include or not each citation.

### 2.3. Evaluation of the performance of GPT models to screen citations for inclusion in systematic reviews

For each systematic review, we compared the recommendations from GPT models (considered as the index test) to rule-out citations based on title and abstract and the original decisions from the authors of the systematic review, at title and abstract level, (considered as the reference standard) by calculating the sensitivity and specificity with 95% confidence intervals. As the systematic reviews were not in the training data set from GPT 3.5-Turbo, we can consider that decisions from GPT models and from authors were completely independent.

As human reviewers can make errors during title and abstract screening, their decisions can be considered as an imperfect reference standard. We therefore considered two reference standards. Reference standard 1 was the original decisions from authors, after title and abstract screening. Reference standard 2 involved the reappraisal of all discrepancies between recommendations from GPT models and original decisions from authors (i.e., “over inclusion”, “missed citation because of a screening error” or “correct original decision”) ^27,28^.

Pooled estimates for sensitivity and specificity across all reviews were obtained using the bivariate model from Reitsma et al. for meta-analysis of diagnostic accuracy studies. This model preserves the two-dimensional nature of the underlying data (i.e., sensitivity and specificity) as compared to models relying solely on the diagnostic odds ratio ^29^. Results were presented by plotting estimates of the paired observed sensitivities and specificities, for each review, and a summary receiver operating characteristic (SROC) curve obtained from the bivariate model aforementioned.

We performed a sensitivity analysis where the performance of GPT models was tested only in citations not used to fine-tune models (i.e., from 80 to 90% citations per review).

### 2.4. Avoidable workload when using of GPT models to screen citations for inclusion in systematic reviews

To assess the workload arising from title and abstract screening, one reviewer (FA) searched PubMed via MEDLINE for all systematic reviews (with or without meta-analyses) evaluating the effectiveness of a pharmacological or biological therapeutic or preventive intervention in humans published between July 1, 2022 and December 31, 2022 (**Appendix 2**). The reviewer excluded studies focused solely on natural medicine, animal studies, diagnostic studies and protocols. The search was performed on October 24, 2023. Among identified reviews, we randomly selected 100 and assessed: 1) the number of citations obtained from the electronic database searches, after deduplication; 2) the number of citations kept after title and abstract screening; 3) the number of citations kept after full-text assessment; 4) the number of reviewers involved in the screening; and 5) whether an automated tool was used to accelerate screening.

We estimated the time required to perform screening using the estimates reported in the Cochrane Handbook, which considers that the results of a database search can be screened at the rate of 60–120 per hour ^26^. Workload was multiplied by the number of reviewers performing screening independently. Whenever the number of reviewers was not reported, we made the conservative hypothesis that only a single reviewer was involved. If the review reported the use of machine learning tool to accelerate screening, we considered 40% average time savings ^9^. To obtain estimates over a one-year period, we hypothesized that 1) our sample of 100 reviews was representative of the identified reviews; and 2) the number of reviews published per year was twice the number of reviews identified over a 6-months period. For example, we considered that the number of citations to be screened over one year, was twice average number of citations observed in the sample multiplied by the number of systematic reviews and meta-analyses identified in the search. To obtain person-years estimates, we arbitrarily considered that a human reviewer was working 8 hours a day for 200 days per year.

Time required for the use of GPT models for title and abstract screening involved 1) human screening for 20% of the number of citations retrieved from electronic searches and 2) a conservative and arbitrary quantity of 8 hours of work to adapt and fine-tune the prompt.

Avoidable workload was obtained by subtracting the time required for human screening and the time required to use GPT models in two hypothetical scenarios: 1) in systematic reviews and meta-analyses where two reviewers performed screening on title and abstract, one reviewer was replaced by the use of a GPT model; and 2) human screening on title and abstract was limited to citations which were not “ruled out” by a GPT model.

## 3. Results

### 3.1. Systematic reviews

The five systematic reviews used in this study identified a total number of 22,666 citations from electronic searches on the Epistemonikos COVID-19 LOVE platform (n=2), the iSearch COVID-19 portfolio (n=1), MEDLINE (n=3), Cochrane Central Register of Controlled Trials (n=2), EMBASE (n=2), Web of Science (n=1), and LILACS (Latin American and Caribbean Health Science Information database) (n=1). After screening of title and abstracts, 1,485 (6.5%) citations were included (ranging from 1.2% to 35%). After full-text screening, 432 (1.9%) citations were included (ranging from 0.1% to 4%) (**Table 1**).

**Table 1.**
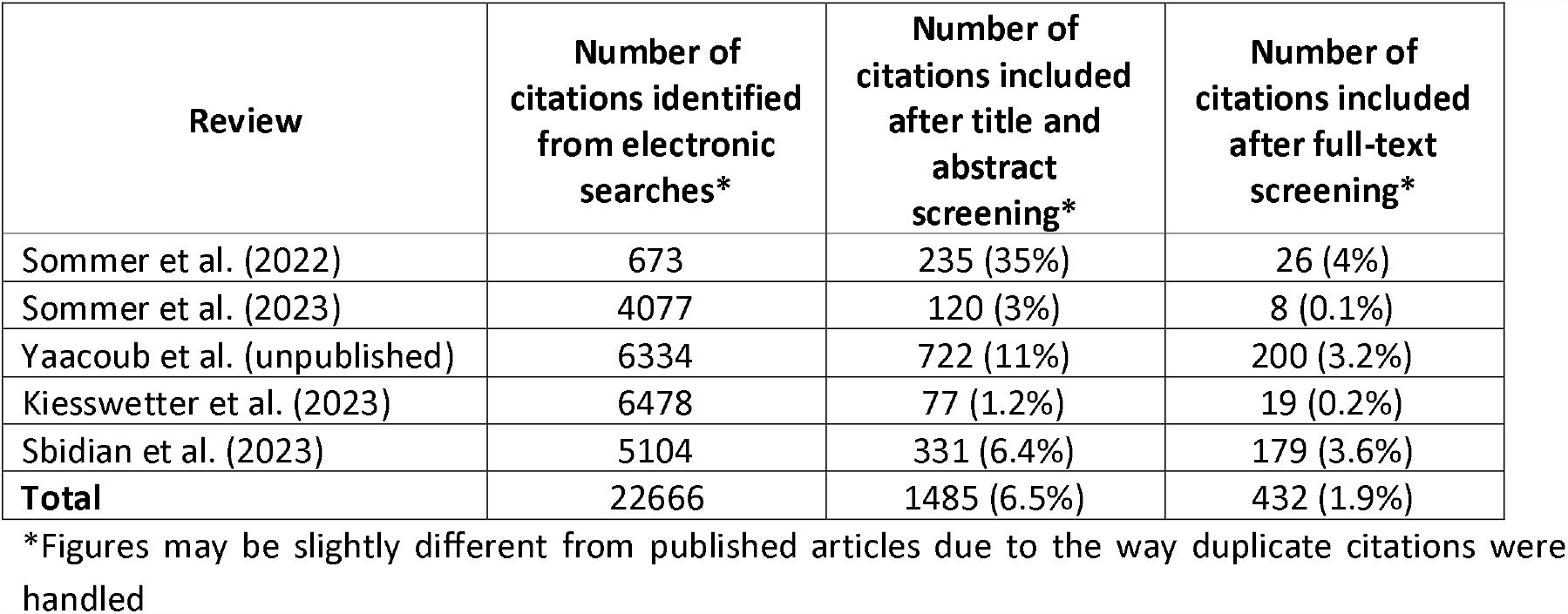
Characteristics of included reviews.

### 3.2. Evaluation of the performance of GPT models to screen citations for inclusion in systematic reviews

Considering the reference standard 1 (i.e. original decisions from authors), sensitivity of GPT models to rule out citations during title and abstract screening for systematic reviews ranged from 81.3% (95% CI 77.1 to 84.9) for the review on the efficacy of primary treatment of confirmed COVID-19 in outpatient settings; to 99.2% (95% CI 99.0 to 99.4) for the review on dairy intake on intermediate disease markers in adults ^22,24^. Specificity ranged from 9.1% (95%CI 6.4 to 12.7) for the review on dairy intake on intermediate disease markers in adults to 56.6% (95%CI 50.8 to 62.2) for the review on the efficacy of primary treatment of confirmed COVID-19 in outpatient settings ^22,24^ (**Table 2**).

**Table 2.** Sensitivity and specificity of GPT models to rule out citations during title and abstract screening. *Reference standard 2 accounts for the imperfect nature of reference standard 1: all discrepancies between decisions from authors and from AI were reassessed by the authors of reviews. **Obtained by using a bivariate model

| Review | Reference standard 1 |  | Reference standard 2* |  |
| --- | --- | --- | --- | --- |
|  | Sensitivity [95%CI] | Specificity [95%CI] | Sensitivity [95%CI] | Specificity [95%CI] |
| Sommer et al. (2022) | 81.3<br>[77.1 – 84.9] | 56.6<br>[50.8 – 62.2] | 87.5<br>[83.9 – 90.5] | 66.0<br>[60.3 – 71.2] |
| Sommer et al. (2023) | 98.1<br>[97.6 – 98.5] | 38.6<br>[30.2 – 47.8] | 99.1<br>[98.7 – 99.3] | 38.6<br>[30.2 – 47.8] |
| Yaacoub et al. (unpublished) | 92.0<br>[91.3 – 92.7] | 52.2<br>[47.8 – 56.6] | 94.6<br>[94.0 – 95.1] | 57.7<br>[53.5 – 62.2] |
| Kiesswetter et al. (2023) | 99.2<br>[99.0 – 99.4] | 9.1<br>[6.4 – 12.7] | 99.6<br>[99.5 – 99.8] | 9.7<br>[7.0 – 13.4] |
| Sbidian et al. (2023) | 91.6<br>[90.7 – 92.3] | 24.8<br>[21.7-28.3] | 92.5<br>[91.7 – 93.2] | 31.7<br>[28.3-35.4] |
| **Pooled statistic\*\*** | **95.3**<br>**[86.2 – 98.5]** | **33.0**<br>**[16.5 – 55.1]** | **97.1**<br>**[89.6 – 99.2]** | **37.7**<br>**[18.4 – 61.9]** |

Considering the reference standard 2 (i.e., decisions from authors and reappraisal of all discrepancies), 97 citations (median 27 (IQR 2 to 27) per review) were excluded on title and abstract by human reviewers despite being eligible for full-text review (i.e., screening errors) and identified by GPT models. On the contrary, GPT models excluded 187 citations (median 19 (IQR 6 to 64) per review) that were considered eligible by human reviewers and included after full-text review (**Appendix 4**). Using the reference standard 2, sensitivity ranged from 87.5% (95%CI 83.9 to 90.5) to 99.6% (95%CI 99.5 to 99.8). Specificity ranged from 9.7% (95%CI 7.0 to 13.4) to 66.0% (95%CI 60.3 to 71.2) (**Table 2**).

Figure 1 summarizes information, displaying the pooled estimates for sensitivity of 97.1% (95% CI 89.6 to 99.2) and for specificity of 37.7% (95% CI 18.4 to 61.9), using reference standard 2. The figure also shows the coherence between sensitivity and specificity across the different reviews, hinting that the performance estimated was not driven by outlier results. Similar information is displayed in **Appendix 3** for the reference standard 1.

**Figure 1:**
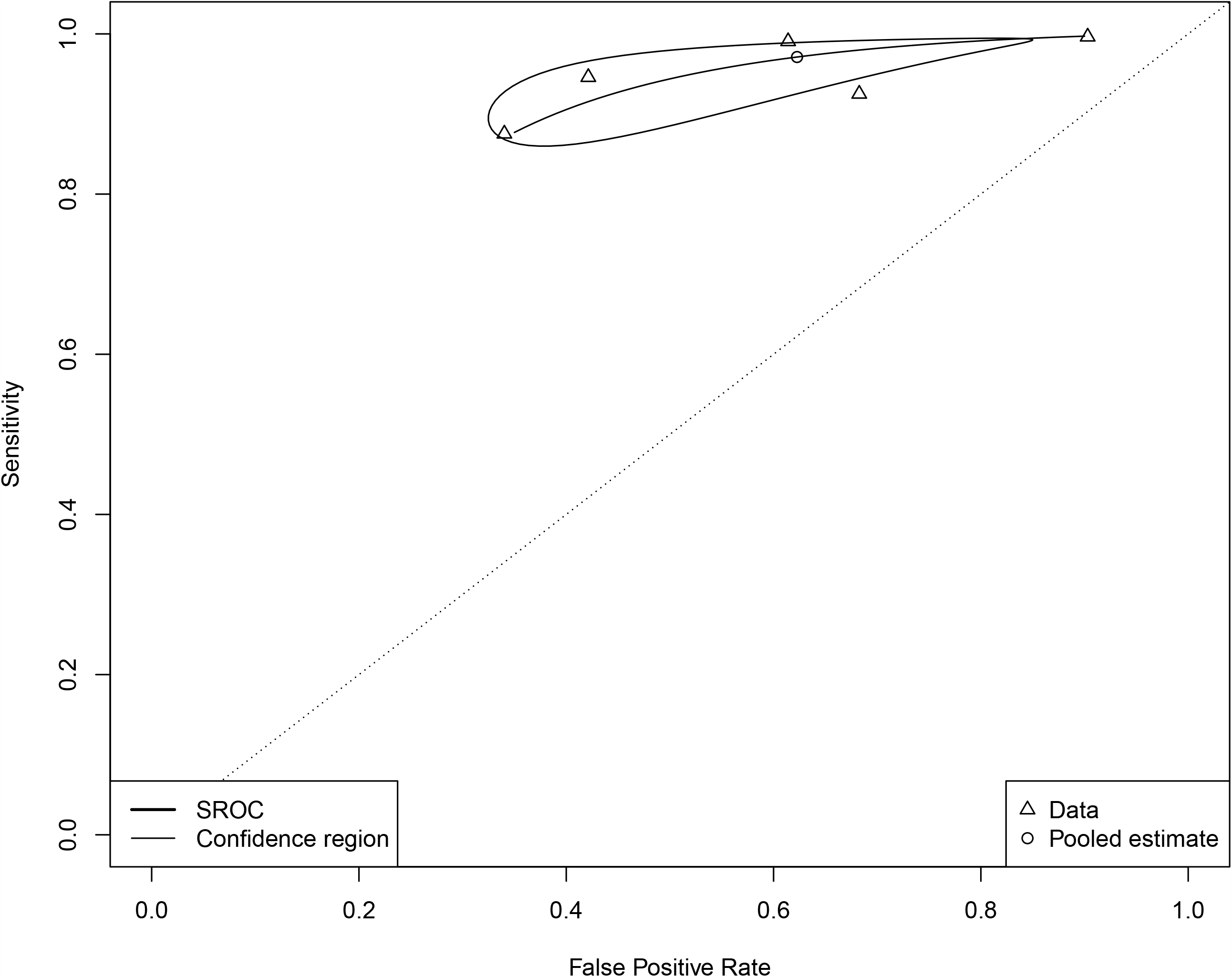
Bivariate summary estimates of sensitivity and specificity for the ability of GPT models to screen for title and abstract in the five systematic reviews. We used reference standard 2 which accounts for imperfect nature of human decisions by reanalysis of all discrepancies by the authors of reviews.

Performance was unchanged in a sensitivity analysis where the performance of GPT models was tested only in citations not used to fine-tune models (i.e., from 80 to 90% citations per review) (**Appendix 5**).

### 3.3. Avoidable workload when using GPT models to screen citations for inclusion in systematic reviews

In total, we identified 2507 systematic reviews evaluating the effectiveness of therapeutic or preventive pharmacological interventions in humans, published between 01/07/2022 and 31/12/2022. In a random sample of 100 studies (**Appendix 3**), the median number of citations 1) identified from electronic search was 662 (interquartile range (IQR 202 to 1740), 2) included after title and abstract screening was 36 (IQR 23 to 74) (median proportion of citations retrieved from electronic searches: 8%, IQR 3 to 21), and 3) included after full-text screening was 13 (IQR 8 to 22) (median proportion: 2.5%, IQR 0.8 to 6.1). The cumulated time to perform title and abstract screening for the 100 reviews was 4208 to 8417 person-hours (i.e., 2.6 to 5.3 person-years of work). By extrapolating these results to 2507*2 systematic reviews evaluating the effectiveness of therapeutic or preventive pharmacological interventions published in 2022, we estimate the workload of title and abstract screening to 211,013 to 422,025 person-hours (i.e., 132 to 264 persons-year of work), every year.

Using GPT models to replace one reviewer in reviews where title and abstract screening was performed in double and independently would reduce workload from 11 to 21% of the workload of title and abstract screening and would save a cumulated time of 24,136 to 88,385 persons hours (15 to 55 person-years of work) per year, as compared to human screening alone. At review level, average time saved by review ranged from 4 to 17 hours per review.

Limiting human screening to citations which were not “ruled out” by GPT models, independently would reduce workload from 50 to 65% of the workload of title and abstract screening and save a cumulated time of 106,268 to 276,053 persons hours (i.e.,66 to 172 person-years of work) per year, as compared to human screening alone. At review level, average time saved by review ranged from 5 to 55 hours per review.

## 4. Discussion

AI systems based on LLMs provide highly sensitive and low to moderate specific recommendations to rule out citations during title and abstract screening in systematic reviews and meta-analyses, without specific training. Using these models as a “triage tool”, used before human screening, could reduce human workload up to 65% and save up to 275,000 person-hours of work per year for systematic reviews evaluating the effectiveness of therapeutic or prophylactic pharmacological interventions on humans and referenced in Medline.

The use of LLM based AI systems to perform title and abstract screening in systematic reviews differ from existing automated tools which learn from the researchers’ decisions to prioritize citations by relevance ^9,30^. First, the tool functions as a zero-shot classifier: decision to exclude a citation is provided without prior training, using solely a prompt based on the PICOS elements reported in the protocol of the review ^31^. As a result, the tool’s performance is not dependent on the proportion of citations that will be included by researchers, nor is it subject to the lack of appropriate stopping criteria faced by prioritization tools ^32^. Second, GPT models use instructions in human language and can therefore be used widely without specific training nor configuration. Third, they function independently from humans, without fatigue. Finally, newer generative AI are multimodal, accept larger inputs, including the upload of documents, potentially suggesting that an AI could bypass title and abstract screening and use all data available to make a recommendation.

In this study, sensitivity of GPT models to rule out citations from title and abstract was high (>87%). However, specificity was more heterogeneous and varied from 10% to 50%. Two factors may have played a role in this result. First, specificity seemed to be associated with the complexity of inclusion and exclusion criteria in our study. For example, in the review evaluating the effect of dairy intake on cardiometabolic health ^24^, decisions often required expert knowledge beyond simply understanding the text (e.g., the protocol described eligible interventions as *“Non-bovine milk and dairy products (e.g. from sheep, goats, buffalos, camels), milk/ protein isolates (e.g. whey or casein), capsules, fortified dairy products (e.g. fortified with Vitamin D, plant sterols/ stanols, prebiotics, probiotics or omega-3 fatty acids) and fermented milk products with additional microbiota strains added (beyond those naturally occurring) will be excluded*). Second, performance also depended on human researchers’ tendency and habits to “over include” at title and abstract screening. This was highlighted by the reassessment of discrepancies between human and AI decisions which could improve specificity up to 10%. Despite low specificity, GPT models are still useful to rule out citations and reduce human workload during the conduct of reviews. For example, in the aforementioned review, pre-screening of citations by GPT models could rule out 6148/6478 (95%) references, helping human reviewers focus on the 330 remaining citations.

Despite high sensitivity, GPT models excluded some citations that had been finally included in reviews after full texts. In particular, we chose to include “outcomes” in the prompt so as to balance sensitivity and specificity, despite some researchers advising not to select on title and abstract based on this information, because some outcomes may not be reported in the abstract or because of selective outcome reporting bias. Furthermore, some exclusion of eligible citations came from the unreliable ability of GPT models to provide expected answers to prompts (“YES” or “NO”) and because the program was designed to capture these answers. This led to a reduction in sensitivity. This highlights that these models are not yet ready to replace humans but may rather be used to complement human assessment (e.g., serving as a second or third reviewer). In all, we show that use of LLM based models can reduce the workload of title and abstract screening by human researchers. For some reviews, a low loss in precision may be acceptable, in particular when considering the time gained, e.g., when conducting rapid reviews for urgent decision-making. Furthermore, use of GPT models might allow to broaden the scope of the searches (e.g., increasing time ranges, relaxing search terms, adding another electronic database in the search, or searching in clinical trial registries) thereby increasing overall comprehensiveness of study inclusion. In particular, the ability of AI-based systems to drastically reduce the human workload is of critical importance with the rise of network meta-analyses and living network meta-analyses ^33^, which typically involve larger literature streams than traditional reviews; as well as rapid reviews, which require the timely analysis of these literature streams ^34^.

We chose to use GPT 3.5-Turbo models to ensure that none of the systematic reviews were in the training data set of the model and that no information about the reference standard results were available to inform the index test. GPT-4 models have shown better performance in most tasks as compared to GPT 3.5-Turbo models and the performance estimates we show here are likely to improve quickly in the next months or years.

Another issue is the reproducibility and transparency of results from GPT models: for example, researchers have shown important differences in models’ performance over a one-month interval ^35^. In this study, highest performance was observed with the first reviews assessed. While we cannot ascertain that time and modification of the models was responsible for the change in performance, this affects the reproducibility of results in reviews. Potential solutions may involve the development and use of “fixed” LLM systems for research tasks.

Our study has several limitations. First, estimates of sensitivity and performance used five reviews from researchers from Cochrane Austria, Germany and France, which involved experienced reviewers and larger searches. Results may therefore differ in other settings. Second, estimation of the avoidable workload relied on multiple hypotheses which may have impacted the final results. As we used conservative hypotheses and given the magnitude of the avoidable workload of evidence synthesis, we believe that AI-based LLM should be considered as a “triage tool”, used before human screening, when performing systematic reviews. Third, it is known that performance of LLM is driven by the prompt used. In this study, we chose a standardized prompting style across all reviews, always following the same structure, and incorporating the PICOS extracted from review protocols. While this approach facilitates the use of our results in different contexts, it is possible that fine-tuned prompts may improve the performance of GPT models in specific reviews.

AI systems based on LLMs provide highly sensitive and low to moderately specific recommendations to include citations during title and abstract screening, in systematic reviews and meta-analyses. Using these models to rule out citations before human screening could significantly reduce human work in evidence synthesis.

## Data Availability

Data from reviews is available upon request to original authors.

## 5. Acknowledgments

We would like to thank Emilie Sbidian, Laurence Le Cleach and Kathrin Grummich for their help in obtaining the unannotated databases.

## 6. Data sharing statement

Unannotated data from reviews are available to academic research teams from contacting original authors of reviews.

## 7. Contributorship statement

Generated the idea: VTT, Conceived and designed the experiments: VTT, GG, JM and PR. Collected data: GG, SY, IB, LS, JS, IS, FA, SA, JM. Analyzed data: VTT, GG, SY, LS, JS, IS, FA and SA; Wrote the first draft of the manuscript: VTT. Contributed to the writing of the manuscript: all authors; ICMJE criteria for authorship read and met: all authors. Agree with manuscript results and conclusions: all authors. VTT is the guarantor. He had full access to the data in the study and take responsibility for the integrity of the data and the accuracy of the data analysis.

## References

1. Donnelly CA, Boyd I, Campbell P, et al. Four principles to make evidence synthesis more useful for policy. Nature. Jun 2018;558(7710):361–364. doi:10.1038/d41586-018-05414-4

2. National Institute for Health and Care Excellence. NICE health technology evaluations: the manual. 2022.

3. Bornmann L, Haunschild R, Mutz R. Growth rates of modern science: a latent piecewise growth curve approach to model publication numbers from established and new literature databases. Humanities and Social Sciences Communications. 2021;8(224)

4. Wang Z, Nayfeh T, Tetzlaff J, O’Blenis P, Murad MH. Error rates of human reviewers during abstract screening in systematic reviews. PloS one. 2020;15(1):e0227742. doi:10.1371/journal.pone.0227742

5. Waffenschmidt S, Knelangen M, Sieben W, Bühn S, Pieper D. Single screening versus conventional double screening for study selection in systematic reviews: a methodological systematic review. BMC medical research methodology. Jun 28 2019;19(1):132. doi:10.1186/s12874-019-0782-0

6. O’Hearn K, MacDonald C, Tsampalieros A, et al. Evaluating the relationship between citation set size, team size and screening methods used in systematic reviews: a cross-sectional study. BMC medical research methodology. Jul 8 2021;21(1):142. doi:10.1186/s12874-021-01335-5

7. Allen IE, Olkin I. Estimating time to conduct a meta-analysis from number of citations retrieved. Jama. Aug 18 1999;282(7):634–5. doi:10.1001/jama.282.7.634

8. Holzinger A. Interactive machine learning for health informatics: when do we need the human-in-the-loop? Brain Inform. jJun 2016;3(2):119–131. doi:10.1007/s40708-016-0042-6

9. Ouzzani M, Hammady H, Fedorowicz Z, Elmagarmid A. Rayyan-a web and mobile app for systematic reviews. Systematic reviews. Dec 5 2016;5(1):210. doi:10.1186/s13643-016-0384-4

10. Norman CR, Leeflang MMG, Porcher R, Névéol A. Measuring the impact of screening automation on meta-analyses of diagnostic test accuracy. Syst Rev. Oct 28 2019;8(1):243. doi:10.1186/s13643-019-1162-x

11. Kilicoglu H, Demner-Fushman D, Rindflesch TC, Wilczynski NL, Haynes RB. Towards automatic recognition of scientifically rigorous clinical research evidence. Journal of the American Medical Informatics Association : JAMIA. Jan-Feb 2009;16(1):25–31. doi:10.1197/jamia.M2996

12. van de Schoot R, de Bruin J, Schram R, et al. An open source machine learning framework for efficient and transparent systematic reviews. Nature Machine Intelligence. 2021;3:125–133.

13. Sarraju A, Bruemmer D, Van Iterson E, Cho L, Rodriguez F, Laffin L. Appropriateness of Cardiovascular Disease Prevention Recommendations Obtained From a Popular Online Chat-Based Artificial Intelligence Model. Jama. Mar 14 2023;329(10):842–844. doi:10.1001/jama.2023.1044

14. Ayers JW, Poliak A, Dredze M, et al. Comparing Physician and Artificial Intelligence Chatbot Responses to Patient Questions Posted to a Public Social Media Forum. JAMA Intern Med. Jun 1 2023;183(6):589–596. doi:10.1001/jamainternmed.2023.1838

15. Levine DM, Tuwani R, Kompa B, et al. The Diagnostic and Triage Accuracy of the GPT-3 Artificial Intelligence Model. medRxiv. 2023;doi:10.1101/2023.01.30.23285067

16. Kung TH, Cheatham M, Medenilla A, et al. Performance of ChatGPT on USMLE: Potential for AI-assisted medical education using large language models. PLOS digital health. Feb 2023;2(2):e0000198. doi:10.1371/journal.pdig.0000198

17. S. W, Liu Y, Xu Y, Zhu C, Zeng M. Want To Reduce Labeling Cost? GPT-3 Can Help. arXiv. 2021;

18. Ding B, Qin C, Liu L, Bing L, Joty S, Li B. Is GPT-3 a Good Data Annotator? arXiv. 2022;doi:10.48550/arXiv.2212.10450

19. Syriani E, David I, Kumar G. Assessing the Ability of ChatGPT to Screen Articles for Systematic Reviews. arXiv. 2023;doi:10.48550/arXiv.2307.06464

20. Guo E, Gupta M, Deng J, Park Y, Paget M, Naugler C. Automated Paper Screening for Clinical Reviews Using Large Language Models arXiv. 2023;doi:10.48550/arXiv.2305.00844

21. Khralsha Q, Put S, Kappenberg J, Warraitch A, Hadfield K. Can large language models replace humans in the systematic review process ? Evaluating GPT-4’s efficacy in screening and extracting data from peer-reviewed and grey literature in multiple languages. ArXiv. 2023;doi:https://arxiv.org/abs/2310.17526

22. Sommer I, Dobrescu A, Ledinger D, et al. Outpatient Treatment of Confirmed COVID-19: A Living, Rapid Review for the American College of Physicians. Annals of internal medicine. Jan 2023;176(1):92–104. doi:10.7326/m22-2202

23. Sommer I, Ledinger D, Thaler K, et al. Outpatient Treatment of Confirmed COVID-19: A Living, Rapid Evidence Review for the American College of Physicians (Version 2). Annals of internal medicine. Sep 19 2023;doi:10.7326/m23-1626

24. Kiesswetter E, Stadelmaier J, Petropoulou M, et al. Effects of Dairy Intake on Markers of Cardiometabolic Health in Adults: A Systematic Review with Network Meta-Analysis. Adv Nutr. May 2023;14(3):438–450. doi:10.1016/j.advnut.2023.03.004

25. Sbidian E, Chaimani A, Guelimi R, et al. Systemic pharmacological treatments for chronic plaque psoriasis: a network meta-analysis. The Cochrane database of systematic reviews. Jul 12 2023;7(7):Cd011535. doi:10.1002/14651858.CD011535.pub6

26. Higgins J, Thomas J, Chandler J, et al. Cochrane Handbook for Systematic Reviews of Interventions version 6.4. Cochrane Collaboration; 2023.

27. Reitsma JB, Rutjes AW, Khan KS, Coomarasamy A, Bossuyt PM. A review of solutions for diagnostic accuracy studies with an imperfect or missing reference standard. J Clin Epidemiol. Aug 2009;62(8):797–806. doi:10.1016/j.jclinepi.2009.02.005

28. Whiting P, Rutjes AW, Reitsma JB, Glas AS, Bossuyt PM, Kleijnen J. Sources of variation and bias in studies of diagnostic accuracy: a systematic review. Ann Intern Med. Feb 3 2004;140(3):189–202. doi:10.7326/0003-4819-140-3-200402030-00010

29. Reitsma JB, Glas AS, Rutjes AW, Scholten RJ, Bossuyt PM, Zwinderman AH. Bivariate analysis of sensitivity and specificity produces informative summary measures in diagnostic reviews. Journal of clinical epidemiology. Oct 2005;58(10):982–90. doi:10.1016/j.jclinepi.2005.02.022

30. Lerner I, Crequit P, Ravaud P, Atal I. Automatic screening using word embeddings achieved high sensitivity and workload reduction for updating living network meta-analyses. Journal of clinical epidemiology. Apr 2019;108:86–94. doi:10.1016/j.jclinepi.2018.12.001

31. Moreno-Garcia C, Jayne C, Elyan E, Aceves-Martins M. A novel application of machine learning and zero-shot classification methods for automated abstract screening in systematic reviews. Decision Analytics Journal. 2023;6(100162)

32. Callaghan MW, Müller-Hansen F. Statistical stopping criteria for automated screening in systematic reviews. Systematic reviews. Nov 28 2020;9(1):273. doi:10.1186/s13643-020-01521-4

33. Zarin W, Veroniki AA, Nincic V, et al. Characteristics and knowledge synthesis approach for 456 network meta-analyses: a scoping review. BMC medicine. Jan 5 2017;15(1):3. doi:10.1186/s12916-016-0764-6

34. Crequit P, Trinquart L, Yavchitz A, Ravaud P. Wasted research when systematic reviews fail to provide a complete and up-to-date evidence synthesis: the example of lung cancer. BMC medicine. Jan 20 2016;14:8. doi:10.1186/s12916-016-0555-0

35. Chen L, Zaharia M, Zou J. How Is ChatGPT’s Behavior Changing over Time. arXiv. 2023;doi:10.48550/arXiv.2307.09009

