## Appendices for "Sensitivity, specificity and avoidable workload of using a large language models for title and abstract screening in systematic reviews and meta-analyses"

### **Appendix 1. Final GPT 3.5-Turbo « Prompts » for the five systematic reviews**

**For the review on outpatient treatment for confirmed COVID-19 (Sommer I, Ann Intern Med, 2022)**

prompthosp <- "Read the following text and pay close attention to details. Answer only with the words \"YES\" or \"NO\" by using the following algorithm:

If the population of the study is hospitalized patients, answer \"NO\".

If the population of the study is patients in ICU, answer \"NO\".

If the population of the study is severe COVID-19 patients, answer \"NO\".

If the population of the study is outpatients (that is patients outside of hospital or nonhospitalized patients), answer \"YES\"

If it is unclear whether the population is outpatients, answer \"YES\"."

promptrct2 <- "Read the following text and pay close attention to details. Answer only with the words \"YES\" or \"NO\" by using the following algorithm:

If the study is a randomized controlled trial, answer \"YES\".

If the study is an observational study, answer \"NO\".

If the study is a review study, answer \"NO\".

If the text reports the protocol of a randomized trial, without results, answer \"NO\".

If the study has no comparator group, answer \"NO\".

If it is unclear whether the text is a randomized controlled trial, answer \"YES\"."

promptinterv <- "Read the following text and pay close attention to details. Answer only with the words \"YES\" or \"NO\" by using the following algorithm:

If the intervention aims at treating COVID-19, with one of the following drugs: chloroquine or hydroxychloroquine, convalescent plasma, lopinavir and ritonavir, ivermectin, molnupiravir, monoclonal antibodies, nitazoxanide, remdesivir, fluvoxamine, azithromycin, or corticosteroids, answer \"YES\".

If the intervention involves only anticoagulants, antiplatelet therapy or vitamins, answer \"NO\".

If the intervention aims at preventing infection by COVID-19 (pre-exposure or post-exposure prophylaxis), answer \"NO\".

If it is unclear whether the intervention meets the above criteria or not, answer \"YES\"."

promptcontrol <- "Read the following text and pay close attention to details. Answer only with the words \"YES\" or \"NO\" by using the following algorithm:

If the control is Placebo, or Usual care, or a Different dose or duration of the intervention, answer \"YES\".

If the control is an active treatment, different from the intervention, answer \"NO\".

If it is unclear whether the control is a Placebo, or Usual care, or a Different dose or duration of the intervention, answer \"YES\"."

promptoutcome <- "Read the following text and pay close attention to details. Answer only with the words \"YES\" or \"NO\" by using the following algorithm:

If at least one of following outcome has been collected: All-cause mortality, COVID-19 specific mortality, recovery, clinical improvement, Admission to hospital due to COVID-19, Health-related quality of life or adverse events, answer \"YES\".

otherwise, answer \"NO\".

If it is unclear wheter the outcome meets the above criteria, answer \"YES\"."

**For the review on outpatient treatment for confirmed COVID-19 (Sommer I, Ann Intern Med, 2023)**

prompthosp <- "Read the following text and pay close attention to details. Proceed step by step. First, assess the inclusion criteria. Second, answer with the words \"YES\" or \"NO\" by using the following algorithm:\n

1-If the study is on outpatients (patients outside of hospital or nonhospitalized patients) with COVID-19, answer \"YES\".\n

2-If the study is on patients with a disease other than COVID-19, answer \"NO\".\n

3-If the study is on hospitalized patients, answer \"NO\".\n

4-If the study is on patients in ICU, answer \"NO\".\n

5-If the study is on severe COVID-19 patients, answer \"NO\".\n

6-If unclear, answer \"NO\"."

promptrct2 <- "Read the following text and pay close attention to details. Proceed step by step. First, assess the study design. Second, answer with the words \"YES\" or \"NO\" by using the following algorithm:\n

1-If the study is a randomized trial, answer \"YES\".\n

2-If the study is an observational study with more than 5000 patients, answer \"YES\".\n

2-If the study is an observational study with less than 5000 participants, answer \"NO\".\n

4-If the study is a case report, answer \"NO\".\n

5-If the study is a case control study, answer \"NO\".\n

6-If the study is a review or a systematic review, or a meta-analysis or a network meta-analysis, answer \"NO\".\n

7-If the study is a modelling study, answer \"NO\".\n

8-If the study is a pharmacokinetic or pharmacodynamic study, answer \"NO\".\n

9-If the text reports the protocol of a randomized trial, without results, answer \"NO\".\n

10-If the study has no control group, answer \"NO\".\n

11-If unclear, answer \"NO\"."

promptinterv <- "Read the following text and pay close attention to details. Proceed step by step. First, assess the intervention. Second, answer only with the words \"YES\" or \"NO\" by using the following algorithm:\n

1-If the study is evaluating chloroquine or hydroxychloroquine or convalescent plasma or lopinavir + ritonavir or ivermectin or molnupiravir or any monoclonal antibody or nirmatrelvir + ritonavir or nitazoxanide or remdesivir or fluvoxamine or azithromycin or corticosteroids or chlorpheniramine or colchicine or ensitrelvir or favipiravir or oral camostat mesylate or metformin or niclosamide, answer \"YES\".\n

2-If the study is evaluating only anticoagulants, antiplatelet therapy, nutrients or vitamins, answer \"NO\".\n

3-If the study aims at preventing infection by COVID-19 (pre-exposure or post-exposure prophylaxis), answer \"NO\".\n

4-If the study aims at treating long covid (also called post-acute sequelae of COVID-19 or post COVID-19 condition), answer \"NO\".\n

5-If the study aims at treating a disease different from COVID-19 (SARS-COV2 infection), answer \"NO\".\n

6-If unclear, answer \"NO\"."

promptcontrol <- "Read the following text and pay close attention to details. Proceed step by step. First, assess the control. Second, answer with the words \"YES\" or \"NO\" by using the following algorithm:\n

1-If the control is a placebo, or usual care, or a different dose or duration of the intervention, answer \"YES\".\n

3-If the control is an active treatment, different from the intervention, answer \"NO\".\n

4-If unclear, answer \"NO\"."

promptoutcome <- "Read the following text and pay close attention to details.Proceed step by step. First, Assess the study outcomes. Second answer with the words \"YES\" or \"NO\" by using the following algorithm:\n

1- If one of following outcome has been collected: All-cause mortality, COVID-19 specific mortality, recovery, clinical improvement, Admission to hospital due to COVID-19, Health-related quality of life or adverse events, answer \"YES\".\n

2- Otherwise, answer \"NO\".\n

3- If unclear, answer \"NO\"."

promptperiod <- "Read the following text and pay close attention to details.Proceed step by step. First, assess the time when the study recruited patients. Second answer with the words \"YES\" or \"NO\" by using the following algorithm:\n

1-If the text contains the word omicron, answer \"YES\".\n

2-If the study enrolled patients in 2022, answer \"YES\".\n

3-If the study enrolled patients before 2022 (for example, in 2020 or in 2021), answer \"NO\".\n

4- If unclear, answer \"NO\"."

**For the methodological review on the epidemiology and reporting characteristics of non-randomized studies of pharmacologic treatment (Yaacoub, unpublished)**

promptrct <- "Read the following text and pay close attention to details. Proceed step by step. First, assess the study design. Second, answer only with the words \"YES\" or \"NO\" by using the following algorithm:

1-If the study is a randomized trial, answer \"NO\".

2-If the study is an observational study with two groups, answer \"YES\".

3-If the study is a case series, answer \"NO\".

4-If the study is a case report, answer \"NO\".

5-If the study is a guideline, answer \"NO\".

6-If the study is an interrupted time series, answer \"NO\".

7-If the study is a case control study, answer \"NO\".

8-If the study is a review, a systematic review or a meta-analysis of several studies, answer \"NO\".

9-If the study is a modelling study, answer \"NO\".

10-If the study is a pharmacokinetic or pharmacodynamic study, answer \"NO\".

11-If the text is the protocol of a randomized trial, without results, answer \"NO\".

12-If the text is an editorial or an opinion piece, answer \"NO\".

13-If the study has no comparator group, answer \"NO\".

14- In other cases, answer \"NO\"."

promptinterv <- "Read the following text and pay close attention to details. Proceed step by step. First, assess the treatment. Second, answer only with the words \"YES\" or \"NO\" by using the following algorithm:

1-If the study evaluates a pharmacological treatment (a drug, medication), answer \"YES\".

2-Otherwise, answer \"NO\".

3-If unclear, answer \"NO\"."

**For the review of Dairy Intake on markers of cardiometabolic health in adults (Kiesswette E, Advances in Nutrition, 2023)**

prompthosp <- "Read the following text and pay close attention to details. Proceed step by step. First, assess the inclusion criteria. Second, answer with the words \"YES\" or \"NO\" by using the following algorithm:\n

1-If the study is on healthy adults, answer \"YES\".\n

2-If the study includes children and adolescents, answer \"NO\".\n

3-If the study includes pregnant women, answer \"NO\".\n

4-If the study includes unhealthy patients or patients with chronic diseases such as diabetes, hypertension or cancer, answer \"NO\".\n

5-If unclear, answer \"NO\"."

promptrct2 <- "Read the following text and pay close attention to details. Proceed step by step. First, assess the study design. Second, answer with the words \"YES\" or \"NO\" by using the following algorithm:\n

1-If the study is a randomized trial, answer \"YES\".\n

2-If the study uses words such as random allocation, answer \"YES\".\n

3-If the study is an observational study, answer \"NO\".\n

4-If the study is a cohort study, answer \"NO\".\n

5-If the study is a case report, answer \"NO\".\n

6-If the study is a case control study, answer \"NO\".\n

7-If the study is a review or a systematic review, or a meta-analysis or a network meta-analysis, answer \"NO\".\n

8-If the study is a modelling study, answer \"NO\".\n

9-If the study is a pharmacokinetic or pharmacodynamic study, answer \"NO\".\n

10-If the text reports the protocol of a randomized trial, without results, answer \"NO\".\n

11-Otherwise, answer \"NO\".

12-If unclear, answer \"NO\"."

promptinterv <- "Read the following text and pay close attention to details. Proceed step by step. First, assess the intervention. Second, answer only with the words \"YES\" or \"NO\" by using the following algorithm:\n

1-If the intervention is based on dairy intake, and if NONE of the following (2 to 10) criteria is true, answer \"YES\".\n

2-If the intervention is NOT a dairy product (milk, cheese or yogurt), answer \"NO\".\n

3-If the intervention states the use of non-bovine dairy products, answer \"NO\".\n

4-If the intervention involves only protein isolates (for example whey or casein), answer \"NO\".\n

5-If the intervention fortifies or enriches dairy products with sterols or stanols, answer \"NO\".\n

6-If the intervention fortifies or enriches dairy products with Vitamin D or omega-3 fatty acids, answer \"NO\".\n

7-If the intervention fortifies or enriches dairy products with probiotics or prebiotics, answer \"NO\".\n

8-If the intervention fermented milk products with additional microbiota strains added, answer \"NO\".\n

9-Otherwise, answer \"NO\".

10-If unclear, answer \"NO\"."

promptcontrol <- "Read the following text and pay close attention to details. Proceed step by step. First, assess the control. Second, answer with the words \"YES\" or \"NO\" by using the following algorithm:\n

1- If the control is a dairy product, answer \"YES\".\n

2- If the control is usual diet, answer \"YES\".\n

3- Otherwise answer \"NO\".\n

4- If unclear, answer \"NO\"."

promptoutcome <- "Read the following text and pay close attention to details.Proceed step by step. First, Assess the variables collected. Second answer with the words \"YES\" or \"NO\" by using the following algorithm:\n

1- If body weight has been collected, answer \"YES\".\n

2- If fat mass has been collected, answer \"YES\".\n

3- If waist circumference has been collected, answer \"YES\".\n

4- If LDL-cholesterol has been collected, answer \"YES\".\n

5- If HDL-cholesterol has been collected, answer \"YES\".\n

6- If triacylglycerol has been collected, answer \"YES\".\n

7- If fasting glucose has been collected, answer \"YES\".\n

8- If glycosylated haemoglobin (HbA1c) has been collected, answer \"YES\".\n

9- If systolic blood pressure has been collected, answer \"YES\".\n

10- Otherwise (none of the above data have been collected), answer \"NO\".\n

11- If unclear, answer \"NO\"."

**For the Cochrane review on systemic pharmacological treatments for chronic plaque psoriasis (Sbidian, Cochrane Database Syst Rev, 2022)**

prompthosp <- "Read the following text and pay close attention to details. Proceed step by step. First, assess the study population. Second, answer with the words \"YES\" or \"NO\" by using the following algorithm:\n

1-If the study includes adult patients with moderate to severe plaque psoriasis, answer \"YES\".\n

2-If the study includes adult patients with psoriatic arthritis with skin lesions considered moderate to severe psoriasis, answer \"YES\".\n

2-If the study includes adult patients with psoriatic arthritis but skin lesions are not described, answer \"NO\".\n

3-If the study includes adult patients with mild psoriasis, answer \"NO\".\n

4-If unclear, answer \"NO\"."

promptrct2 <- "Read the following text and pay close attention to details. Proceed step by step. First, assess the study design. Second, answer with the words \"YES\" or \"NO\" by using the following algorithm:\n

1-If the study is a randomized trial (random allocation of patients), answer \"YES\".\n

2-If the study is a phase 1 randomized trial, answer \"NO\".\n

3-If the study is a cross-over trial, answer \"NO\".\n

4-If the study is an observational study , answer \"NO\".\n

5-If the study is a case report, answer \"NO\".\n

6-If the study is a case control study, answer \"NO\".\n

7-If the study is a review or a systematic review, or a meta-analysis or a network meta-analysis, answer \"NO\".\n

8-If the study is a modelling study, answer \"NO\".\n

9-If the study is a pharmacokinetic or pharmacodynamic study, answer \"NO\".\n

10-If the text reports the protocol of a randomized trial and does not include numerical results, answer \"NO\".\n

11-If the text describes a study that has not started, answer \"NO\".\n

12-If the study has no control group, answer \"NO\".\n

13-If unclear, answer \"NO\"."

promptinterv <- "Read the following text and pay close attention to details. Proceed step by step. First, assess the intervention, that is the drug under study. Second, answer only with the words \"YES\" or \"NO\" by using the following algorithm:\n

1-If the intervention is FAEs, answer \"YES\".\n

2-If the intervention is Acitretin, answer \"YES\".\n

3-If the intervention is Ciclosporin, answer \"YES\".\n

4-If the intervention is Methotrexate, answer \"YES\".\n

5-If the intervention is Apremilast, answer \"YES\".\n

6-If the intervention is Tofacitinib, answer \"YES\".\n

7-If the intervention is Ponesimod, answer \"YES\".\n

8-If the intervention is Infliximab, answer \"YES\".\n

9-If the intervention is Etanercept, answer \"YES\".\n

10-If the intervention is Adalimumab, answer \"YES\".\n

11-If the intervention is Certolizumab, answer \"YES\".\n

12-If the intervention is Ustekinumab, answer \"YES\".\n

13-If the intervention is Secukinumab, answer \"YES\".\n

14-If the intervention is Brodalumab, answer \"YES\".\n

15-If the intervention is Ixekizumab, answer \"YES\".\n

16-If the intervention is Tildrakizumab, answer \"YES\".\n

17-If the intervention is Guselkumab, answer \"YES\".\n

18-If the intervention is Itolizumab, answer \"YES\".\n

19-If the intervention is Alefacept, answer \"YES\".\n

20-If the intervention drug is a combination of several molecules, answer \"NO\".\n

21-Otherwise (drug is NOT in proposition 1 to 19), answer \"NO\".\n

22-If unclear, answer \"NO\"."

promptcontrol <- "Read the following text and pay close attention to details. Proceed step by step. First, assess the control. Second, answer with the words \"YES\" or \"NO\" by using the following algorithm:\n

1-If the control is FAEs, answer \"YES\".\n

2-If the control is Acitretin, answer \"YES\".\n

3-If the control is Ciclosporin, answer \"YES\".\n

4-If the control is Methotrexate, answer \"YES\".\n

5-If the control is Apremilast, answer \"YES\".\n

6-If the control is Tofacitinib, answer \"YES\".\n

7-If the control is Ponesimod, answer \"YES\".\n

8-If the control is Infliximab, answer \"YES\".\n

9-If the control is Etanercept, answer \"YES\".\n

10-If the control is Adalimumab, answer \"YES\".\n

11-If the control is Certolizumab, answer \"YES\".\n

12-If the control is Ustekinumab, answer \"YES\".\n

13-If the control is Secukinumab, answer \"YES\".\n

14-If the control is Brodalumab, answer \"YES\".\n

15-If the control is Ixekizumab, answer \"YES\".\n

16-If the control is Tildrakizumab, answer \"YES\".\n

17-If the control is Guselkumab, answer \"YES\".\n

18-If the control is Itolizumab, answer \"YES\".\n

19-If the control is Alefacept, answer \"YES\".\n

20-If the control is standard psoriasis treatment such as topical treatment or phototherapy, answer \"YES\".\n

22-If the control is the same treatment as the intervention, but with, for example, a different dose, answer \"NO\".\n

21-If unclear, answer \"NO\"."

promptoutcome <- "Read the following text and pay close attention to details. Proceed step by step. First, Assess the outcomes (also called endpoints) measured in the study. Second answer with the words \"YES\" or \"NO\" by using the following algorithm:\n

1- If the text mentions the Psoriasis Area and Severity Index score, also called PASI, answer \"YES\".\n

2- If outcomes only include cellular or biological data, answer \"NO\".\n

3- Otherwise, answer \"NO\".\n

4- If unclear, answer \"NO\"."

### **Appendix 2. Search strategy to identify the systematic reviews and meta-analyses evaluating the effectiveness of a pharmacological or biological therapeutic or prophylactic intervention on humans published between July 1, 2022 and December 31, 2022 & references of the 100 studies selected**

**PubMed search**

(("Meta-Analysis" [Publication Type]) OR "Meta-Analysis as Topic"[Mesh]) NOT (Prevalence[ti] OR Exposure[ti] OR Life style[ti] OR Risk factor[ti] OR Exercise[ti] OR Food[ti] OR Yoga[ti])

Dates : 2022/07/01 - 2022/12/31

**References of the sample of 100 studies used to evaluate the workload of research**

1.	Wang XH, Wang ZQ, Mu ZY, Zhu LP, Zhong CF, Guo S. The efficacy and safety of immune checkpoint inhibitors in metastatic castration-resistant prostate cancer: A systematic review and meta-analysis. Medicine. 2022 Aug 5;101(31):e29715.

2.	Yingyan Z, Huasheng L, Jingyao Y, Xiaohong HE, Lili P, Xue LI, et al. Effectiveness and safety of tripterygium glycosides tablet for lupus nephritis: a systematic review and Meta-analysis. J Tradit Chin Med. 2022 Oct;42(5):671–80.

3.	Guo X, Liu C, Huang Y. Efficacy and Safety of Vitamin D Adjuvant Therapy for Ulcerative Colitis: A Meta-Analysis. Comput Math Methods Med. 2022;2022:6836942.

4.	Michael HU, Youbi E, Ohadoma SC, Ramlall S, Oosthuizen F, Polyakova M. A Meta-Analytic Review of the Effect of Antiretroviral Therapy on Neurocognitive Outcomes in Adults Living with HIV-1 in Low-and Middle-Income Countries. Neuropsychology Review 2021 32:4 [Internet]. 2021 Nov 10 [cited 2023 Oct 11];32(4):828–54. Available from: https://link.springer.com/article/10.1007/s11065-021-09527-y

5.	Diallo A, Carlos-Bolumbu M, Renard PE, Galtier F. Larger effect size in composite kidney outcomes than in major cardiovascular events associated with sodium-glucose cotransporter-2 (SGLT2) inhibitors compared with glucagon-like peptide-1 receptor agonists (GLP-1RAs): A pooled analysis of type 2 diabetes trials. Diabetes Obes Metab. 2023 Jan;25(1):166–76.

6.	Yu CL, Chou PY, Liang CS, Tu YK, Chi CC. Interventions for molluscum contagiosum: A systematic review and network meta-analysis with normalized entropy assessment. J Am Acad Dermatol [Internet]. 2023 Feb 1 [cited 2023 Aug 28];88(2):508–10. Available from: http://www.jaad.org/article/S0190962222023490/fulltext

7.	Ismaila AS, Haeussler K, Czira A, Tongbram V, Malmenäs M, Agarwal J, et al. Comparative Efficacy of Umeclidinium/Vilanterol Versus Other Bronchodilators for the Treatment of Chronic Obstructive Pulmonary Disease: A Network Meta-Analysis. Adv Ther [Internet]. 2022 Nov 1 [cited 2023 Aug 31];39(11):4961–5010. Available from: https://pubmed.ncbi.nlm.nih.gov/35857184/

8.	Maida M, Ventimiglia M, Facciorusso A, Vitello A, Sinagra E, Marasco G. Effectiveness and safety of 1-L PEG-ASC versus other bowel preparations for colonoscopy: A meta-analysis of nine randomized clinical trials. Dig Liver Dis. 2023 Aug;55(8):1010–8.

9.	Fakhrolmobasheri M, Abhari AP, Manshaee B, Heidarpour M, Shafie D, Mohammadbeigi E, et al. Effect of sodium-glucose cotransporter 2 inhibitors on insulin resistance; a systematic review and meta-analysis. Acta Diabetol. 2023 Feb;60(2):191–202.

10.	van Dijk EHC, Feenstra HMA, Bjerager J, Grauslund J, Boon CJF, Subhi Y. Comparative efficacy of treatments for chronic central serous chorioretinopathy: A systematic review with network meta-analyses. Acta Ophthalmol. 2023 Mar;101(2):140–59.

11.	Tian Z, Li Y, Xie Y, Yang Y, Xu J. Efficacy and safety of tacrolimus combined with corticosteroids in patients with idiopathic membranous nephropathy: a systematic review and meta-analysis of randomized controlled trials. Int Urol Nephrol. 2022 Oct;54(10):2555–66.

12.	Gulia S, Kannan S, Ghosh J, Rath S, Maheshwari A, Gupta S. Maintenance therapy with a poly(ADP-ribose) polymerase inhibitor in patients with newly diagnosed advanced epithelial ovarian cancer: individual patient data and trial-level meta-analysis. ESMO Open. 2022 Oct;7(5):100558.

13.	Zhu A, Kuznia S, Boakye D, Schöttker B, Brenner H. Vitamin D-Binding Protein, Bioavailable, and Free 25(OH)D, and Mortality: A Systematic Review and Meta-Analysis. Nutrients. 2022 Sep 20;14(19).

14.	Albuquerque TR de, Macedo LFR, Delmondes G de A, Rolim Neto ML, Almeida TM, Uchida RR, et al. Evidence for the beneficial effect of ketamine in the treatment of patients with post-traumatic stress disorder: A systematic review and meta-analysis. J Cereb Blood Flow Metab. 2022 Dec;42(12):2175–87.

15.	Dwivedi P, Patel TK, Bajpai V, Singh Y, Tripathi A, Kishore S. Efficacy and safety of intranasal ketamine compared with intranasal dexmedetomidine as a premedication before general anesthesia in pediatric patients: a systematic review and meta-analysis of randomized controlled trials. Can J Anaesth. 2022 Nov;69(11):1405–18.

16.	Lijuan D, Lu Y, Liping W, Yaya S, Xiaoyun L, Xiangrong LI. Effectiveness of redcore lotion in patients with vulvovaginal candidiasis: a systematic review and Meta-analysis. J Tradit Chin Med. 2022 Aug;42(4):487–92.

17.	Garg A, Rout A, Farhan S, Waxman S, Giustino G, Tayal R, et al. Dual antiplatelet therapy duration after percutaneous coronary intervention using drug eluting stents in high bleeding risk patients: A systematic review and meta-analysis. Am Heart J. 2022 Aug;250:1–10.

18.	Tao S, Huang J, Xiao J, Ke G, Fu P. Cardio-selective versus non-selective β-blockers for cardiovascular events and mortality in long-term dialysis patients: A systematic review and meta-analysis. PLoS One. 2022;17(12):e0279171.

19.	Wang Y, Yu L, Ma D, Lu L, Liu B, Liu Z, et al. Efficacy and safety of filgotinib in patients with rheumatoid arthritis and inadequate response to disease-modifying antirheumatic drugs (DMARDs): A meta-analysis of randomized controlled trials. ARP rheumatology. 2022 Oct 1;1(ARP Rheumatology, no3 2022):230–43.

20.	Chai S, Zhang R, Zhang Y, Carr RD, Zheng Y, Rajpathak S, et al. Influence of dipeptidyl peptidase-4 inhibitors on glycemic variability in patients with type 2 diabetes: A meta-analysis of randomized controlled trials. Front Endocrinol (Lausanne). 2022;13:935039.

21.	Yang CW, Chen RD, Zhu QR, Han SJ, Kuang MJ. Efficacy of umbilical cord mesenchymal stromal cells for COVID-19: A systematic review and meta-analysis. Front Immunol. 2022;13:923286.

22.	Hasegawa D, Lee YI, Prasitlumkum N, Chopra L, Nishida K, Smith RL, et al. Premorbid angiotensin converting enzyme inhibitors or angiotensin II receptor blockers in patients with sepsis. Am J Emerg Med. 2022 Dec;62:69–77.

23.	Tsukamoto S, Morita R, Yamada T, Urate S, Azushima K, Uneda K, et al. Cardiovascular and kidney outcomes of combination therapy with sodium-glucose cotransporter-2 inhibitors and mineralocorticoid receptor antagonists in patients with type 2 diabetes and chronic kidney disease: A systematic review and network meta-analysis. Diabetes Res Clin Pract. 2022 Dec;194:110161.

24.	Botta C, Gigliotta E, Paiva B, Anselmo R, Santoro M, Otero PR, et al. Network meta-analysis of randomized trials in multiple myeloma: Efficacy and safety in frontline therapy for patients not eligible for transplant. Hematol Oncol [Internet]. 2022 Dec 1 [cited 2023 Aug 28];40(5):987–98. Available from: https://onlinelibrary.wiley.com/doi/full/10.1002/hon.3041

25.	Orenday-Barraza JM, Cavagnaro MJ, Avila MJ, Strouse IM, Farhadi DS, Dowell A, et al. Is the routine use of systemic antibiotics after spine surgery warranted? A systematic review and meta-analysis. European Spine Journal [Internet]. 2022 Oct 1 [cited 2023 Oct 18];31(10):2481–92. Available from: https://link.springer.com/article/10.1007/s00586-022-07294-9

26.	Yan H, Shi J, Li X, Dai Y, Wu Y, Zhang J, et al. Oral gonadotropin-releasing hormone antagonists for treating endometriosis-associated pain: a systematic review and network meta-analysis. Fertil Steril. 2022 Dec;118(6):1102–16.

27.	Chen Q, Liang Y, Yan J, Du Y, Li M, Chen Z, et al. Efficacy and safety of non-steroidal mineralocorticoid receptor antagonists for renal outcomes: A systematic review and meta-analysis. Diabetes Res Clin Pract. 2023 Jan;195:110210.

28.	Chen L, Ni Y, Wu X, Chen G. Probiotics for the prevention of atopic dermatitis in infants from different geographic regions: a systematic review and Meta-analysis. J Dermatolog Treat. 2022 Nov;33(7):2931–9.

29.	Gui T, Li H, Zhu F, Wang Q, Zhou X, Xue Q. Different dosage regimens of erenumab for the treatment of migraine: A systematic review and meta-analysis of the efficacy and safety of randomized controlled trials. Headache. 2022 Nov;62(10):1281–92.

30.	Li Y, Du Y, Xue C, Wu P, Du N, Zhu G, et al. Efficacy and safety of anti-PD-1/PD-L1 therapy in the treatment of advanced colorectal cancer: a meta-analysis. BMC Gastroenterol. 2022 Oct 10;22(1):431.

31.	Farhadian N, Farhadian M, Zamanian MH, Taghadosi M, Vaziri S. Sotrovimab therapy in solid organ transplant recipients with mild to moderate COVID-19: a systematic review and meta-analysis. https://doi.org/101080/0892397320222160733 [Internet]. 2022 [cited 2023 Sep 18]; Available from: https://www.tandfonline.com/doi/abs/10.1080/08923973.2022.2160733

32.	Zhou B, Xia H, Yang L, Wang S, Sun G. The Effect of Lycium Barbarum Polysaccharide on the Glucose and Lipid Metabolism: A Systematic Review and Meta-Analysis. Journal of the American Nutrition Association. 2022 Aug;41(6):618–26.

33.	Lin GX, Chen CM, Zhu MT, Zheng L. The Safety and Effectiveness of Tranexamic Acid in Lumbar Interbody Fusion Surgery: An Updated Meta-analysis of Randomized Controlled Trials. World Neurosurg. 2022 Oct 1;166:198–211.

34.	Keum N, Chen QY, Lee DH, Manson JE, Giovannucci E. Vitamin D supplementation and total cancer incidence and mortality by daily vs. infrequent large-bolus dosing strategies: a meta-analysis of randomised controlled trials. Br J Cancer. 2022 Sep;127(5):872–8.

35.	Dehghani F, Abdollahi S, Shidfar F, Clark CCT, Soltani S. Probiotics supplementation and brain-derived neurotrophic factor (BDNF): a systematic review and meta-analysis of randomized controlled trials. Nutr Neurosci [Internet]. 2023 Oct 3 [cited 2023 Oct 9];26(10):942–52. Available from: https://www.tandfonline.com/doi/abs/10.1080/1028415X.2022.2110664

36.	Issa PP, Hussein M, Omar M, Munshi R, Attia AS, Buti Y, et al. Cardiovascular Health by Graves’ Disease Management Modality - Surgery Versus Radioactive Iodine Versus Antithyroid Medications: A Network Meta-Analysis. J Surg Res [Internet]. 2023 Mar 1 [cited 2023 Aug 30];283:266–73. Available from: https://pubmed.ncbi.nlm.nih.gov/36423475/

37.	Bao Y, Ma Z, Yuan M, Wang Y, Men Y, Hui Z. Comparison of different neoadjuvant treatments for resectable locoregional esophageal cancer: A systematic review and network meta-analysis. Thorac Cancer [Internet]. 2022 Sep 1 [cited 2023 Aug 28];13(17):2515–23. Available from: https://onlinelibrary.wiley.com/doi/full/10.1111/1759-7714.14588

38.	Liu M, Yuan Y, Qiao Y, Tang Y, Sui X, Yin P, et al. The effectiveness of immunomodulatory therapies for patients with repeated implantation failure: a systematic review and network meta-analysis. Scientific Reports 2022 12:1 [Internet]. 2022 Nov 1 [cited 2023 Aug 28];12(1):1–12. Available from: https://www.nature.com/articles/s41598-022-21014-9

39.	Katsaras DN, Katsaras GN, Chatziravdeli VI, Papavasileiou GN, Touloupaki M, Mitsiakos G, et al. Comparative safety and efficacy of paracetamol versus non-steroidal anti-inflammatory agents in neonates with patent ductus arteriosus: A systematic review and meta-analysis of randomized controlled trials. Br J Clin Pharmacol [Internet]. 2022 Jul 1 [cited 2023 Oct 20];88(7):3078–100. Available from: https://onlinelibrary.wiley.com/doi/full/10.1111/bcp.15291

40.	Idaikkadar P, Georgiou A, Skene S, Michael A. Non-Surgical Management of Malignant Bowel Obstruction in Advanced Ovarian Cancer patients: A Systematic Review and Meta-Analysis. Am J Hosp Palliat Care. 2022 Jul;39(7):838–46.

41.	Abdallah M, Brown L, Provenza J, Tariq R, Gowda S, Singal AK. Safety and efficacy of dyslipidemia treatment in NAFLD patients: a meta-analysis of randomized controlled trials. Ann Hepatol [Internet]. 2022 Nov 1 [cited 2023 Sep 1];27(6). Available from: https://pubmed.ncbi.nlm.nih.gov/35781090/

42.	Lai CC, Wang YH, Chen KH, Chen CH, Wang CY. The Clinical Efficacy and Safety of Anti-Viral Agents for Non-Hospitalized Patients with COVID-19: A Systematic Review and Network Meta-Analysis of Randomized Controlled Trials. Viruses [Internet]. 2022 Aug 1 [cited 2023 Aug 29];14(8). Available from: /pmc/articles/PMC9415971/

43.	Su H, Lu Y, Ma C, Li H, Su X. Impact of atorvastatin on erectile dysfunction: A meta-analysis and systematic review. Andrologia [Internet]. 2022 Jul 1 [cited 2023 Oct 20];54(6):e14408. Available from: https://onlinelibrary.wiley.com/doi/full/10.1111/and.14408

44.	Zhang Y, Jiang L, Wang J, Wang T, Chien C, Huang W, et al. Network meta-analysis on the effects of finerenone versus SGLT2 inhibitors and GLP-1 receptor agonists on cardiovascular and renal outcomes in patients with type 2 diabetes mellitus and chronic kidney disease. Cardiovasc Diabetol [Internet]. 2022 Dec 1 [cited 2023 Sep 1];21(1). Available from: /pmc/articles/PMC9637313/

45.	Park DY, Wang P, An S, Grimshaw AA, Frampton J, Ohman EM, et al. Shortening the duration of dual antiplatelet therapy after percutaneous coronary intervention for acute coronary syndrome: A systematic review and meta-analysis. Am Heart J. 2022 Sep 1;251:101–14.

46.	Shi H, Su X, Yan B, Li C, Wang L. Effects of oral alkali drug therapy on clinical outcomes in pre-dialysis chronic kidney disease patients: a systematic review and meta-analysis. Ren Fail [Internet]. 2022 Dec 31 [cited 2023 Oct 6];44(1):106–15. Available from: https://www.tandfonline.com/doi/abs/10.1080/0886022X.2021.2023023

47.	Wang Z, Li W, Lyu Z, Yang L, Wang S, Wang P, et al. Effects of probiotic/prebiotic/synbiotic supplementation on blood glucose profiles: a systematic review and meta-analysis of randomized controlled trials. Public Health. 2022 Sep;210:149–59.

48.	Liabsuetrakul T, Yamamoto Y, Kongkamol C, Ota E, Mori R, Noma H. Medications for preventing hypertensive disorders in high-risk pregnant women: a systematic review and network meta-analysis. Syst Rev [Internet]. 2022 Dec 1 [cited 2023 Aug 29];11(1):1–17. Available from: https://systematicreviewsjournal.biomedcentral.com/articles/10.1186/s13643-022-01978-5

49.	Kato A, Horita N, Namkoong H, Nomura E, Masuhara N, Kaneko T, et al. Prophylactic antibiotics for postcataract surgery endophthalmitis: a systematic review and network meta-analysis of 6.8 million eyes. Sci Rep [Internet]. 2022 Dec 1 [cited 2023 Aug 31];12(1). Available from: https://pubmed.ncbi.nlm.nih.gov/36258003/

50.	Rafizadeh R, Danilewitz M, Bousman CA, Mathew N, White RF, Bahji A, et al. Effects of clozapine treatment on the improvement of substance use disorders other than nicotine in individuals with schizophrenia spectrum disorders: A systematic review and meta-analysis. J Psychopharmacol [Internet]. 2023 Feb 1 [cited 2023 Oct 16];37(2):135. Available from: /pmc/articles/PMC9912304/

51.	Hayashi M, Kano K, Kuroda N, Yamamoto N, Shiroshita A, Kataoka Y. Comparative efficacy of sedation or analgesia methods for reduction of anterior shoulder dislocation: A systematic review and network meta-analysis. Academic Emergency Medicine [Internet]. 2022 Oct 1 [cited 2023 Aug 28];29(10):1160–71. Available from: https://onlinelibrary.wiley.com/doi/full/10.1111/acem.14568

52.	Geng A, Pei T, Zhao X, Jia B. A Meta-analysis of Xiaoyin Granules Combined with Acitretin Capsule in the Treatment of Psoriasis Vulgaris. Comput Math Methods Med [Internet]. 2022 [cited 2023 Oct 10];2022. Available from: /pmc/articles/PMC9348960/

53.	Li ACW, Dong C, Tay ST, Ananthakrishnan A, Ma KSK. Vedolizumab for acute gastrointestinal graft-versus-host disease: A systematic review and meta-analysis. Front Immunol. 2022;13:1025350.

54.	Ehsan M, Rehman AU, Athar F, Mustafa B, Javed H, Cheema HA, et al. Benvitimod for the treatment of psoriasis: A systematic review and meta-analysis of randomized controlled trials. Dermatol Ther. 2022 Dec;35(12):e15957.

55.	Berti A, Alsawas M, Jawaid T, Prokop LJ, Lee JM, Jeong GH, et al. Induction and maintenance of remission with mycophenolate mofetil in ANCA-associated vasculitis: a systematic review and meta-analysis. Nephrol Dial Transplant [Internet]. 2022 Nov 1 [cited 2023 Sep 18];37(11):2190–200. Available from: https://pubmed.ncbi.nlm.nih.gov/34910216/

56.	Haikun W, Na W, Dan S. Efficacy and safety of different doses of baricitinib for rheumatoid arthritis: A Bayesian network meta-analysis. Medicine [Internet]. 2022 Sep 23 [cited 2023 Aug 30];101(38):E30676. Available from: https://pubmed.ncbi.nlm.nih.gov/36197174/

57.	Zeng L, Deng Y, Yang K, Chen J, He Q, Chen H. Safety and efficacy of fecal microbiota transplantation for autoimmune diseases and autoinflammatory diseases: A systematic review and meta-analysis. Front Immunol [Internet]. 2022 Sep 30 [cited 2023 Sep 4];13. Available from: https://pubmed.ncbi.nlm.nih.gov/36248877/

58.	Li X, Sui C, Xia X, Chen X. Efficacy and Safety of Botulinum Toxin Type A for Treatment of Glabellar Lines: A Network Meta-Analysis of Randomized Controlled Trials. Aesthetic Plast Surg [Internet]. 2023 Feb 1 [cited 2023 Aug 28];47(1):365–77. Available from: https://link.springer.com/article/10.1007/s00266-022-03018-y

59.	Luo C, Wu G, Huang X, Ding Y, Huang Y, Song Q, et al. Myeloablative conditioning regimens in adult patients with acute myeloid leukemia undergoing allogeneic hematopoietic stem cell transplantation in complete remission: a systematic review and network meta-analysis. Bone Marrow Transplant. 2023 Feb;58(2):175–85.

60.	Simadibrata DM, Syam AF, Lee YY. A comparison of efficacy and safety of potassium-competitive acid blocker and proton pump inhibitor in gastric acid-related diseases: A systematic review and meta-analysis. J Gastroenterol Hepatol [Internet]. 2022 Dec 1 [cited 2023 Sep 23];37(12):2217–28. Available from: https://pubmed.ncbi.nlm.nih.gov/36181401/

61.	Ke Y, Wang J, Wang W, Guo S, Dai M, Wu L, et al. Antithrombotic strategies after transcatheter aortic valve implantation: A systematic review and network meta-analysis of randomized controlled trials. Int J Cardiol [Internet]. 2022 Sep 1 [cited 2023 Sep 1];362:139–46. Available from: https://pubmed.ncbi.nlm.nih.gov/35654173/

62.	Mori Y, Duru OK, Tuttle KR, Fukuma S, Taura D, Harada N, et al. Sodium-Glucose Cotransporter 2 Inhibitors and New-onset Type 2 Diabetes in Adults With Prediabetes: Systematic Review and Meta-analysis of Randomized Controlled Trials. J Clin Endocrinol Metab. 2022 Dec 17;108(1):221–31.

63.	Holgersson J, Ceric A, Sethi N, Nielsen N, Jakobsen JC. Fever therapy in febrile adults: systematic review with meta-analyses and trial sequential analyses. BMJ. 2022 Jul 12;378:e069620.

64.	Kneißl S, Stallhofer J, Schlattmann P, Stallmach A. Disease recurrence in patients with Crohn’s disease after biologic therapy or surgery: a meta-analysis. Int J Colorectal Dis. 2022 Oct;37(10):2185–95.

65.	Teoh L, McCullough M, Taing MW. Efficacy of oxycodone for postoperative dental pain: A systematic review and meta-analysis. J Dent. 2022 Oct;125:104254.

66.	Kishi T, Sakuma K, Hatano M, Iwata N. N-acetylcysteine for schizophrenia: A systematic review and meta-analysis. Psychiatry Clin Neurosci [Internet]. 2023 Feb 1 [cited 2023 Oct 16];77(2):119–21. Available from: https://onlinelibrary.wiley.com/doi/full/10.1111/pcn.13502

67.	Setayesh L, Pourreza S, Zeinali Khosroshahi M, Asbaghi O, Bagheri R, Rezaei Kelishadi M, et al. The effects of guar gum supplementation on lipid profile in adults: a GRADE-assessed systematic review, meta-regression and dose-response meta-analysis of randomised placebo-controlled trials. Br J Nutr. 2023 May 28;129(10):1703–13.

68.	Long T, Lin JT, Lin MH, Wu QL, Lai JM, Li SZ, et al. Comparative efficiency and safety of insulin degludec/aspart with insulin glargine in type 2 diabetes: a meta-analysis of randomized controlled trials. Endocr J [Internet]. 2022 [cited 2023 Sep 13];69(8):959–69. Available from: https://pubmed.ncbi.nlm.nih.gov/35431280/

69.	An S, Kim K, Kim MH, Jung JH, Kim Y. Perioperative Probiotics Application for Preventing Postoperative Complications in Patients with Colorectal Cancer: A Systematic Review and Meta-Analysis. Medicina (Kaunas). 2022 Nov 14;58(11).

70.	Dai Q, Wang Y, Liao M, Chen H. Efficacy and safety of CDK4/6 inhibitors combined with endocrine therapy versus endocrine therapy alone in hormone receptor-positive, HER2-negative, advanced breast cancer: a systematic review and meta-analysis. Ann Palliat Med [Internet]. 2022 Dec 1 [cited 2023 Oct 6];11(12):3727–42. Available from: https://apm.amegroups.org/article/view/107142/html

71.	Wang J, Liao B, Wang C, Zhong O, Lei X, Yang Y. Effects of Antioxidant Supplementation on Metabolic Disorders in Obese Patients from Randomized Clinical Controls: A Meta-Analysis and Systematic Review. Oxid Med Cell Longev. 2022;2022:7255413.

72.	Bai Z, Wang L, Wang R, Zou M, Méndez-Sánchez N, Romeiro FG, et al. Use of human albumin infusion in cirrhotic patients: a systematic review and meta-analysis of randomized controlled trials. Hepatol Int [Internet]. 2022 Dec 1 [cited 2023 Sep 15];16(6):1468–83. Available from: https://pubmed.ncbi.nlm.nih.gov/36048318/

73.	Ou Y, Feng M, Hu B, Dong Y. The impact of alfentanil supplementation on the sedation of bronchoscopy: A meta-analysis of randomized controlled trials. Medicine [Internet]. 2022 Aug 8 [cited 2023 Oct 4];101(31):E27401. Available from: /pmc/articles/PMC9351902/

74.	Karunakaran M, Kaur R, Ismail S, Cherukuru S, Jonnada PK, Senadhipan B, et al. Post-hepatectomy venous thromboembolism: a systematic review with meta-analysis exploring the role of pharmacological thromboprophylaxis. Langenbecks Arch Surg [Internet]. 2022 Dec 1 [cited 2023 Oct 9];407(8):3221–33. Available from: https://link.springer.com/article/10.1007/s00423-022-02610-9

75.	Masetti R, Muratore E, Gori D, Prete A, Locatelli F. Allogeneic hematopoietic stem cell transplantation for pediatric acute myeloid leukemia in first complete remission: a meta-analysis. Ann Hematol [Internet]. 2022 Nov 1 [cited 2023 Sep 11];101(11):2497–506. Available from: https://link.springer.com/article/10.1007/s00277-022-04965-x

76.	Jalili C, Talebi S, Mehrabani S, Bagheri R, Wong A, Amirian P, et al. Effects of camelina oil supplementation on lipid profile and glycemic control: a systematic review and dose‒response meta-analysis of randomized clinical trials. Lipids Health Dis. 2022 Dec 7;21(1):132.

77.	Vajdi M, Musazadeh V, Karimi A, Heidari H, Tarrahi MJ, Askari G. Effects of Chromium Supplementation on Lipid Profile: an Umbrella of Systematic Review and Meta-analysis. Biol Trace Elem Res [Internet]. 2023 Aug 1 [cited 2023 Oct 19];201(8):3658–69. Available from: https://link.springer.com/article/10.1007/s12011-022-03474-2

78.	Ghasemi F, Navab F, Rouhani MH, Amini P, Shokri-Mashhadi N. The effect of lutein and Zeaxanthine on dyslipidemia: A meta-analysis study. Prostaglandins Other Lipid Mediat. 2023 Feb 1;164:106691.

79.	Abdelazeem B, Awad AK, Manasrah N, Elbadawy MA, Ahmad S, Savarapu P, et al. The Effect of Vasopressin and Methylprednisolone on Return of Spontaneous Circulation in Patients with In-Hospital Cardiac Arrest: A Systematic Review and Meta-analysis of Randomized Controlled Trials. American Journal of Cardiovascular Drugs [Internet]. 2022 Sep 1 [cited 2023 Sep 4];22(5):523–33. Available from: https://link.springer.com/article/10.1007/s40256-022-00522-z

80.	Zhang X, Chen X, Tang Y, Guan X, Deng J, Fan J. Effects of medical plants from Zingiberaceae family on cardiovascular risk factors of type 2 diabetes mellitus: A systematic review and meta-analysis of randomized controlled trials. J Food Biochem [Internet]. 2022 Jul 1 [cited 2023 Oct 20];46(7):e14130. Available from: https://onlinelibrary.wiley.com/doi/full/10.1111/jfbc.14130

81.	Hafkamp FJ, Tio RA, Otterspoor LC, de Greef T, van Steenbergen GJ, van de Ven ART, et al. Optimal effectiveness of heart failure management - an umbrella review of meta-analyses examining the effectiveness of interventions to reduce (re)hospitalizations in heart failure. Heart Fail Rev. 2022 Sep;27(5):1683–748.

82.	Lameire DL, Khalik HA, Phillips M, MacDonald AE, Banfield L, de Sa D, et al. Thromboprophylaxis after knee arthroscopy does not decrease the risk of deep vein thrombosis: a network meta-analysis. Knee Surgery, Sports Traumatology, Arthroscopy [Internet]. 2022 Jul 1 [cited 2023 Aug 31];30(7):2364–76. Available from: https://link.springer.com/article/10.1007/s00167-021-06857-5

83.	Hsieh CF, Tseng PT, Chen TY, Lin PY, Chen YW, Bo-Lin Ho, et al. The association of changes of sleep architecture related to donepezil: A systematic review and meta-analysis. Journal of the Formosan Medical Association. 2022 Aug 1;121(8):1466–77.

84.	Maimaitiming N, Ma X, Wei Y, Cao L, Gao Y, Zhang L. Efficacy and safety of endostar combined with chemoradiotherapy versus chemoradiotherapy alone in locally advanced cervical cancer: A PRISMA-compliant systematic review and meta-analysis. Medicine. 2022 Sep 9;101(36):e30170.

85.	Frey HA, Stout MJ, Abdelwahab M, Tuuli MG, Woolfolk C, Shamshirsaz AA, et al. Vaginal progesterone for preterm birth prevention in women with arrested preterm labor. J Matern Fetal Neonatal Med. 2022 Dec;35(25):8160–8.

86.	Peng DS, Huang BQ, Ning HT, Zhu XZ. Efficacy and safety of PI3K/Akt/mTOR inhibitors combined with trastuzumab therapy for HER2-positive breast cancer: a meta-analysis. Eur Rev Med Pharmacol Sci. 2022;26(20):7667–78.

87.	Fu H, Guo W, Zhou B, Liu Y, Gao Y, Li M. Efficacy and safety of micronized purified flavonoid fractions for the treatment of postoperative hemorrhoid complications: A systematic review and meta-analysis. Phytomedicine. 2022 Sep 1;104:154244.

88.	Yang J ming, Luo Y, Zhang J hong, Liu Q qin, Zhu Q, Ye H, et al. Effects of WB-EMS and protein supplementation on body composition, physical function, metabolism and inflammatory biomarkers in middle-aged and elderly patients with sarcopenic obesity: A meta-analysis of randomized controlled trials. Exp Gerontol. 2022 Sep 1;166:111886.

89.	Zarezadeh M, Musazadeh V, Ghalichi F, Kavyani Z, Nasernia R, Parang M, et al. Effects of probiotics supplementation on blood pressure: An umbrella meta-analysis of randomized controlled trials. Nutr Metab Cardiovasc Dis. 2023 Feb;33(2):275–86.

90.	Yalle-Vásquez S, Osco-Rosales K, Nieto-Gutierrez W, Benites-Zapata V, Pérez-López FR, Alarcon-Ruiz CA. Vitamin E supplementation improves testosterone, glucose- and lipid-related metabolism in women with polycystic ovary syndrome: a meta-analysis of randomized clinical trials. Gynecol Endocrinol [Internet]. 2022 [cited 2023 Sep 15];38(7):548–57. Available from: https://pubmed.ncbi.nlm.nih.gov/35612360/

91.	Shargian L, Raanani P, Yeshurun M, Gafter-Gvili A, Gurion R. Chimeric antigen receptor T-cell therapy is superior to standard of care as second-line therapy for large B-cell lymphoma: A systematic review and meta-analysis. Br J Haematol. 2022 Sep;198(5):838–46.

92.	Marcec R, Dodig VM, Radanovic I, Likic R. Intravenous immunoglobulin (IVIg) therapy in hospitalised adult COVID-19 patients: A systematic review and meta-analysis. Rev Med Virol. 2022 Nov;32(6):e2397.

93.	Zhu L, Chen R, Yang Q, Liu H, Zheng Q, Li L. Modelling an evaluation of the efficacy and safety of gemcitabine combined with platinum in the treatment of non-small cell lung cancer. J Clin Pharm Ther [Internet]. 2022 Jul 1 [cited 2023 Sep 24];47(7):986–94. Available from: https://pubmed.ncbi.nlm.nih.gov/35246996/

94.	Zhao L, Zhu G, Wu L, Xie D. Effects of vitamin D on inflammatory state in patients with chronic kidney disease: A controversial issue. Ther Apher Dial. 2023 Jun;27(3):383–93.

95.	Bhagat S, El-Kafsi J, Samraj K, Mastoridis S. Prophylactic administration of alpha-blockers for the prevention of post-operative urinary retention following inguinal hernia repair: A meta-analysis of randomized control trials. Surgeon. 2023 Aug;21(4):e152–8.

96.	Zhou Q, Han C, Xia Y, Wan F, Yin S, Li Y, et al. Efficacy and safety of 3-n-butylphthalide for the treatment of cognitive impairment: A systematic review and meta-analysis. CNS Neurosci Ther. 2022 Nov;28(11):1706–17.

97.	Putranto R, Harimurti K, Setiati S, Safitri ED, Rizny S, Saldi F, et al. The Effect of Vitamin D Supplementation on Symptoms of Depression in Patients with Type 2 Diabetes Mellitus: A Systematic Review and Meta-Analysis of Randomized Controlled Trials. Acta Med Indones [Internet]. 2023 Jan 10 [cited 2023 Sep 29];54(4):574. Available from: https://www.actamedindones.org/index.php/ijim/article/view/2171

98.	Adam FA, Mohd N, Rani H, Mohd Yusof MYP, Baharin B. A systematic review and meta-analysis on the comparative effectiveness of Salvadora persica - extract mouthwash with chlorhexidine gluconate in periodontal health. J Ethnopharmacol. 2023 Feb 10;302:115863.

99.	Pereira AMG, Pannain GD, Esteves BHG, Bacci ML de L, Rocha MLTLF da, Lopes RGC. Antibiotic prophylaxis in pregnant with premature rupture of ovular membranes: systematic review and meta-analysis. Einstein [Internet]. 2022 [cited 2023 Oct 6];20:eRW0015. Available from: /pmc/articles/PMC9744430/

100.	Zhang Y, Gao YL, Zhang LH. Methylprednisolone and local anesthetic for long-term postherpetic neuralgia: a meta-analysis. J Dermatolog Treat. 2022 Sep;33(6):2723–9.

### **Appendix 3. Bivariate summary estimates of sensitivity and specificity for the ability of GPT models to screen for title and abstract in the five systematic reviews (Reference standard 1: original decisions from authors)**

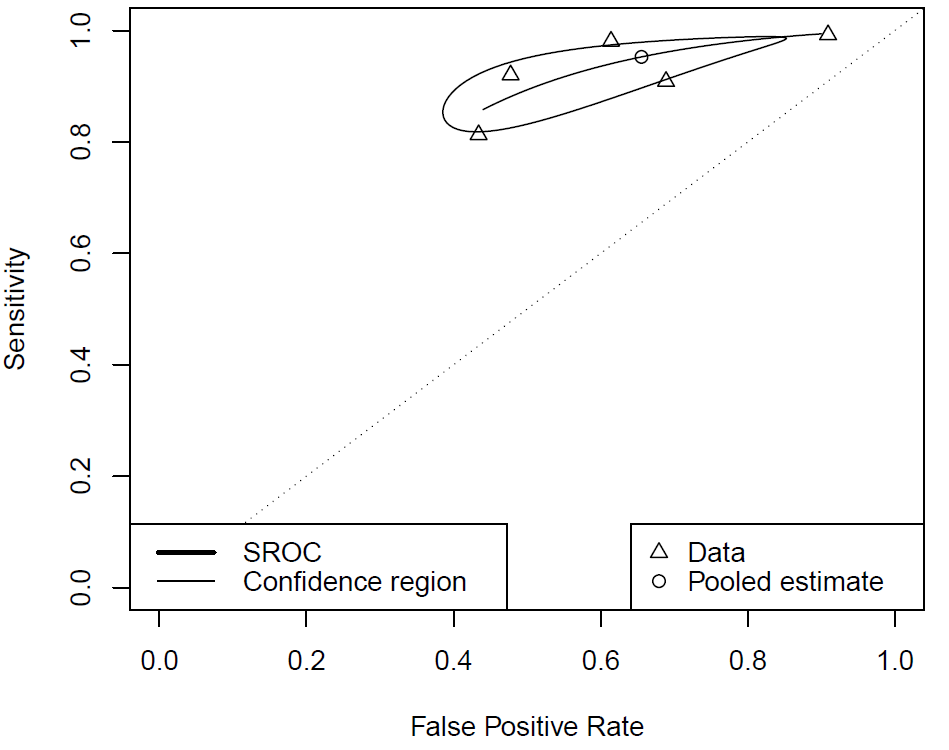

**Appendix 4: References missed by humans and GPT during title and abstract screening**

|  | **Reference standard 2*** | |
| --- | --- | --- |
| **Review** | **Missed by humans during title and abstract screening** | **Missed by GPT during title and abstract screening AND included after full text review** |
| Sommer et al. (2022) | 27 | 0 |
| Sommer et al. (2023) | 0 | 6 |
| Yaacoub et al. (unpublished) | 27 | 98 |
| Kiesswetter et al. (2023) | 2 | 19 |
| Sbidian et al. (2023) | 41 | 64 |

**Appendix 5: Sensitivity analysis of the performance of GPT models to rule out citations during title and abstract screening against reference standard 2, excluding citations used to train the models**

|  | **Reference standard 2*** | |
| --- | --- | --- |
| **Review** | **Sensitivity**  **[95%CI]** | **Specificity**  **[95%CI]** |
| Sommer et al. (2022) | 88.4  [84.6 – 91.3] | 65.5  [59.6 – 71.0] |
| Sommer et al. (2023) | 99.0  [98.6 – 99.2] | 38.1  [29.1 – 48.1] |
| Yaacoub et al. (unpublished) | 94.4  [93.8 – 95.0] | 57.7  [53.1 – 62.3] |
| Kiesswetter et al. (2023) | 99.7  [99.5 – 99.8] | 9.9  [6.8 – 14.1] |
| Sbidian et al. (2023) | 92.4  [91.5 – 93.1] | 31.2  [27.5-35.0] |
| **Pooled statistic**** | 97.2  [89.5 – 99.3] | 37.5  [18.4 – 61.4] |
